# Depth diagnosis of colorectal cancer located on a colorectal fold by quantitative evaluation of a width of colorectal - fold lateral contour using a lateral split-view computed tomographic air-contrast enema image

**DOI:** 10.1101/2024.10.01.24314167

**Authors:** Mitsutoshi Miyasaka, Toshio Muraki, Yusuke Nishimuta, Eiji Oki, Kousei Ishigami, Daisuke Tsurumaru

## Abstract

**Purpose:** To investigate the usefulness of quantitative evaluation of a width of lateral contour on a lateral split-view computed tomographic air-contrast enema (CT enema) image for diagnosing the invasion depth of colorectal cancer (CRC) located on a colorectal fold.

**Methods:** The cases of 22 patients with 22 fold-located CRCs (12 early CRCs and 10 advanced CRCs) who underwent a pretherapeutic computed tomographic colonography were retrospectively examined. T1-stage CRCs were classified into two categories according to the Japanese guideline: T1a-stage, carcinoma invading the superficial submucosa (< 1000 um); and T1b-stage, carcinoma invading the deeper submucosa (≥ 1000 um). The maximum width of a lateral contour of the colorectal fold on which the CRC was located, i.e., the gap distance between the two adjacent haustrations, was calculated from the lateral split-view CT enema image by two gastrointestinal radiologists. These values were compared between the intramucosal / T1a CRCs and the T1b / more deeply invading CRCs. The inter-rater intraclass correlation coefficients were also evaluated for reliability.

**Results:** The maximum widths of a lateral contour of the colorectal fold were significantly higher in the T1b / more deeply invading CRCs than in the intramucosal / T1a CRCs (p<0.0001). The optimum cut-off value of the maximum width of a lateral contour of the colorectal fold for differentiating the former from the latter was 6.0 mm, with a sensitivity and specificity of 100% and 100%, respectively. The inter-rater intraclass correlation coefficient for measurement of a lateral contour of the colorectal fold was 0.958.

**Conclusions:** We demonstrated for the first time that quantitative evaluation of a width of lateral contour using a lateral split-view CT enema image can improve the diagnostic accuracy of the invasion depth for CRCs located on a colorectal fold.

## Background

The invasion depth of colorectal cancer (CRC) lesions is closely related with their risk of metastasis. Surgical resection is the only curative treatment for CRCs with massive submucosal or deeper invasion without distant metastasis, whereas endoscopic resection is the established therapy for intramucosal CRCs or CRCs invading the superficial submucosa (< 1000 um) because they have no risk of metastases [1-4]. Therefore, accurate diagnosis of CRC invasion depth is crucial for determining the appropriate therapeutic strategy in CRC.

Computed tomographic colonography (CTC) is performed worldwide as a preoperative study in CRCs [5-8]. A lateral deformity on CT air-contrast enema (CT enema) or double-contrast barium enema (DCBE) has been shown to be useful in diagnosing the invasion depth of CRCs [9-12]. However, diagnosis of invasion depth based on a lateral deformity may lead to overdiagnosis because the depth of lateral deformity on a CT enema or DCBE image in CRC is understood to depend on both the tumor geometry (mainly with respect to the size of the tumor base) and tumor invasion [13-19]. A CT enema image with cross-sectional multiplanar reconstruction (CS-MPR) images is useful to estimate the depth of lateral deformity based on the tumor geometry in colorectal polyps [20]. Additionally, the quantitative evaluation of lateral deformity on such images improve the accuracy of pretherapeutic diagnosis of the invasion depth of early CRCs [21].

However, when CRC lesions locate on a colorectal fold, it is difficult to estimate quantitatively the depth of lateral deformity based on the tumor geometry on a CT enema image because the lateral deformity on the CT enema image contains not only the lateral deformity caused by both the geometry and invasion of the tumor but also the curved indentation caused by the colorectal fold itself. In clinical practice, though the depth diagnosis of CRC on a colorectal fold using the lateral view of a CT enema or DCBE image is morphologically evaluated by whether width of a colorectal - fold lateral contour is significantly large or not, the evaluation sometimes differs between readers. Accordingly, it is expected that quantitative evaluation of width of a colorectal - fold lateral contour on CT enema images will improve the accuracy of pretherapeutic diagnosis of the invasion depth of CRCs located on a colorectal fold. To our knowledge, there have been no reports on quantitative evaluation of width of a colorectal - fold lateral contour using a CT enema image in the depth diagnosis of CRCs located on a colorectal fold. In the CT enema image, unlike in the DCBE image, it is possible to evaluate CRCs from any view. Therefore, we devised a lateral split-view as a new technique for evaluating the CT enema image and hypothesized that quantitative evaluation of a width of a colorectal - fold lateral contour on CT enema using the lateral split-view would be useful for the depth diagnosis of a CRC located on a colorectal fold. Thus, we conducted the present study to investigate the usefulness of determining the invasion depth of CRCs located on a colorectal fold by quantitative evaluation of widths of a colorectal - fold lateral contour on CT enema, using lateral split-views.

## Methods

### Subjects

The initial patient pool consisted of 142 consecutive patients with CRCs who underwent CTC as part of a pretherapeutic examination at our institution from January 2014 to March 2017. One of the authors reviewed the CTC and endoscopic images in all subjects. Twenty-five patients with 25 CRCs located on a colorectal fold were enrolled and 3 patients with 3 CRCs of pedunculated type located on a colorectal fold were excluded from this study. The final study population included 22 patients (16 males and 6 females) with 22 CRCs located on a colorectal fold. The mean age of the patients was 65 years (range 47–84 years). Eight CRC lesions were resected endoscopically, and the other 14 CRCs were resected by surgery. This retrospective study was approved by the institutional review board of our hospital. The requirement for informed consent was waived. The mean interval between the CTC and the endoscopic or surgical resection was 13 days (range 2–27 days). Data was analyzed between January 9, 2014 and March 31,2017.

### Histology

The macroscopic type of CRCs was classified according to the Paris endoscopic classification [22]. T staging was defined according to the Japanese Classification of Colorectal Carcinoma 9th edition, and early CRCs were classified into three categories: intramucosal carcinoma, high-grade dysplasia in the West; T1a-stage, carcinoma invading the superficial submucosa (< 1000 um); and T1b-stage, carcinoma invading the deeper submucosa (≥ 1000 um) (23).

### CTC Procedure

CTC was performed on the same day of and within 1 hour after a colonoscopy, with bowel preparation using polyethylene glycol. Prior the scanning, colorectal insufflation with carbon dioxide using a CO2 injector (PROTOCO2L; Bracco, Princeton, NJ) was performed. For all patients, the CTC was performed using either a 64-slice multidetector-row computed tomography (MDCT) scanner (Aquilion 64; Canon Medical Systems, Tokyo) or a 320-slice MDCT scanner (Aquilion One; Canon Medical Systems). The scanning parameters were as follows:120 kV, 100–300 mA, beam collimation 1 mm, slice thickness 1 mm, reconstruction interval 1 mm, pitch of 53 or 65. Both supine and prone scans were underwent for all patients. The MDCT data sets were loaded to a workstation (Synapse Vincent, Fujifilm Medical, Tokyo). We reconstructed CT enema images and virtual endoscopic images using the workstation.

### Image analysis

The definition and measurement methods of the width of a colorectal - fold lateral contour on CT enema images are shown in Fig 1. First, we identified CRCs using virtual endoscopic images and CT enema images. Then, we extracted CT enema images in the location of the CRCs using the best distended series. Next, each image was split at the central part of the lesion in the cross-sectional view of the CT enema image, and two images were extracted. Finally, the width of a colorectal - fold lateral contour in the lateral view of each split CT enema image was calculated, and the maximum value of a width of a colorectal - fold lateral contour was defined as the value of a width of a colorectal - fold lateral contour of the CRC located on a colorectal fold. We then compared the maximum width of a colorectal - fold lateral contour between the intramucosal / T1a- stage CRCs and the T1b- stage / more deeply invading CRCs. Additionally, we calculated the width of a normal colorectal - fold lateral contour located closest to the CRCs in a lateral view of a CT enema image. These measurements were performed independently by two gastrointestinal radiologists with 12 and 20 years of experience, respectively, who were given no clinicopathological information except for the lesion locations. The mean value of the maximum width of the colorectal - fold lateral contours of calculated by the two readers was defined as the most probable width of the colorectal - fold lateral contour.

**Fig 1.** Definition and measurement of a width of colorectal - fold lateral contour using lateral split-view CT enema images of a CRC located on a colorectal fold. First, CT enema images in the location of the CRCs were extracted using the best distended series (supine or prone), and then the split images at a central part of the lesion in the cross-sectional view of the CT enema image in the CRCs were extracted. Finally, the value of a width of colorectal - fold lateral contour in the lateral view of each split CT enema image was calculated, and the maximum value of a width of colorectal - fold lateral contour was defined as the value of a width of colorectal - fold lateral contour of the CRC located on a colorectal fold. In this case, the value of a width of colorectal - fold lateral contour calculated using split image 2 was defined as the value of a width of colorectal - fold lateral contour of the CRC located on a fold. L, lesion.

### Statistical analysis

The Student’s t-test was used to assess the differences in the maximum width of a colorectal - fold lateral contour between the intramucosal / T1a-stage and the T1b-stage / more deeply invading CRCs. We also used Receiver operating characteristic (ROC) curves to define the optimum cutoff values for the maximum width of a colorectal - fold lateral contour to differentiate these two groups of CRCs. Additionally, we calculated the intraclass correlation coefficient (ICC) to evaluate the inter-rater reliability of the assessments on the maximum width of a colorectal - fold lateral contour. ICC values of <0.50 were considered to indicate poor reliability, 0.50-0.75 moderate reliability, 0.75-0.90 good reliability and values of >0.90 excellent reliability (24). All statistical analyses were performed using JMP software (JMP version 9.0.2; SAS Institute, Cary, NC). P-values <0.05 were considered significant.

## Results

The clinicopathological characteristics of the CRCs are summarized in Table 1. The average diameter of the CRCs was 18.7 ± 2.0 mm (range 8–38 mm). Twelve lesions were early CRCs and 10 lesions were advanced CRCs (6 lesions were inrtamucosal CRCs, 6 were T1-stage CRCs, 4 were T2-stage CRCs and 6 were T3/T4-stage CRCs). The macroscopic type of CRCs were 3 lesions of type 0-Is, 6 lesions of type 0-Is + IIc, 1 lesion of type 0-IIa, 4 lesions of type 0-IIa + IIc, 3 lesions of type 1, and 5 lesions of type 2. There was no significant difference in the age, sex, tumor size, tumor location or tumor differentiation between the intramucosal / T1a-stage CRC group and the T1b- stage / more deeply invading CRCs group.

**Table 1.** Characteristics of the 22 patients with 22 CRCs

| Clinicopathological factors | Intramucosal / T1a-stage CRCs (n=8) | T1b-stage / more deeply invasive CRCs (n=14) | P value |
| --- | --- | --- | --- |
| Age (years)(range) | 73.1 (65-81) | 66.3 (44-81) | 0.1054 |
| Sex |  |  | 0.8647 |
| Men | 6 | 10 |  |
| Women | 2 | 4 |  |
| Tumor location |  |  | 0.4266 |
| Right – sided colon | 6 | 8 |  |
| Left – sided colon and rectum | 2 | 6 |  |
| Tumor diameter (mm, mean ± SD) | 19.4 (8-38) | 18.8 (11-33) | 0.6982 |
| Tumor differentiation |  |  | 0.1744 |
| Differentiated type | 8 | 11 |  |
| Undifferentiated type | 0 | 3 |  |

Difference in the maximum width of a colorectal - fold lateral contour between the intramucosal / T1a-stage CRCs and T1b-stage / more deeply invading CRCs

There was a significant difference in the maximum width of a colorectal - fold lateral contour between the intramucosal / T1a-stage and T1b-stage / more deeply invading CRCs (3.3 ± 1.4 mm and 12.1 ± 4.6 mm, respectively; p<0.05) (Fig 2). Fig 3 and 4 show representative intramucosal CRC and T1b-stage CRC cases, respectively. The ROC analysis showed that the optimum cutoff value of the maximum width of a colorectal - fold lateral contour for differentiating intramucosal / T1a-stage CRCs from T1b-stage / more deeply invading CRCs was 6 mm, with an AUC of 1. If CRCs with the maximum width of a colorectal - fold lateral contour of >6 mm were interpreted as T1b-stage / more deeply invading CRCs, the sensitivity and specificity for differentiating intramucosal / T1a-stage CRCs from T1b-stage / more deeply invading CRCs were 100% and 100%, respectively. The ICC value for the measurement of a colorectal - fold lateral contour was 0.958, which met the definition for excellent reliability. The mean width of a normal colorectal - fold lateral contour was 2.3 ± 0.6 mm.

**Fig 2.** Comparison of the maximum values of a width of colorectal - fold lateral contour between the intramucosal / T1a-stage CRCs and T1b-stage / more deeply invading CRCs groups. There was a significant difference in the maximum values of a width of colorectal - fold lateral contour between the intramucosal / T1a-stage CRCs and T1b-stage / more deeply invading CRCs (3.3 ± 1.4 mm and 12.1 ± 4.6 mm, respectively; p<0.0001).

**Fig 3.** A 60s-year-old man with intramucosal ascending colon cancer, type 0-IIa. a. Optical endoscopic image shows an elevated lesion located on a colonic fold. b. The values of a width (double arrow) of colonic - fold lateral contour on the lateral split-view CT enema images 1 and 2 in the CRC located on a fold (arrow) were 2.5 mm and 2.4 mm, respectively. Accordingly, the maximum value of a width of colonic - fold lateral contour was diagnosed as 2.5 mm.

**Fig 4.** A 50s-year-old woman with T1b-stage ascending colon cancer, type 0-Is. a. Optical endoscopic image shows an elevated lesion located on a colonic fold. b. The values of a width (double arrow) of colonic - fold lateral contour on the lateral split-view CT enema images 1 and 2 in the CRC located on a colonic fold (arrow) were 2.0 mm and 11.0 mm, respectively. Accordingly, the maximum value of a width of colonic - fold lateral contour was diagnosed as 11.0 mm.

## Discussion

In this study, we reported for the first time on the usefulness of quantitative evaluation of widths of a colorectal - fold lateral contour on CT enema images for the diagnosis of the invasion depth of CRCs located on a colorectal fold. A lateral deformity on a CT enema or DCBE image is one of most important indicators for the diagnosis of invasive CRC, in addition to deep depression, protrusion in depression and fold convergency [9, 12, 14, 25]. In morphologic evaluation of lateral deformity on CT enema or DCBE images in CRCs located on a colorectal fold, CRCs with “a normal width of a colorectal - fold lateral contour” are diagnosed as intramucosal / T1a-stage CRCs and those with “a large width of a colorectal - fold lateral contour” are diagnosed as T1b-stage / more- deeply invading CRCs. However, in our experience, readers morphologically evaluating widths of a colorectal - fold lateral contour on CT enema or DCBE images sometimes have differing opinions as to whether or not a width of a colorectal - fold lateral contour is significantly large. One of the reasons for the disagreement is that lateral views of CT enema or DCB images in CRCs located on a colorectal fold are composed of complex lines— i.e., the indentation of the colorectal fold, tumor surface and tumor base. CT enema imaging is better than DCBE imaging for obtaining a precise and secure lateral view [14, 25]. Furthermore, in the present study, it was possible to simplify these complex lines using lateral split-view CT enema images and to calculate objectively the width of a colorectal - fold lateral contour.

On the other hand, previous studies have reported that lateral deformities based on tumor geometry were detectable in colorectal polyps on DCBE and CT enema images [10, 11, 18]. In CRCs, the depth of the lateral deformity based on tumor geometry depends on the size of the tumor base on a CS-MPR image. Namely, the larger the size of the tumor base is, the larger the value of the depth of lateral deformity based on the tumor geometry will be. It is also necessary to take account of the lateral deformity based on tumor geometry in the depth diagnosis due to lateral deformities in CRCs located on a colorectal fold. However, it is difficult to estimate quantitatively the depth of lateral deformities based on the tumor geometry on CT enema images in CRCs located on a colorectal fold. Nevertheless, in the present study, the diagnostic accuracy of invasion depth for CRCs located on a colorectal fold using the width of a colorectal - fold lateral contour on a CT enema image was very high, even though we could not estimate quantitatively the depth of lateral deformity based on tumor geometry. In the depth diagnosis of CRCs located on a colorectal fold using quantitative evaluation of lateral deformity on a CT enema image, it may not be very important to estimate quantitatively the depth of lateral deformity based on tumor geometry. Endoscopic ultrasound is also useful for determination of the invasion depth for CRCs, with a diagnostic accuracy of approximately 90%, but CRCs located on a colorectal fold are often difficult to image on endoscopic ultrasound [26]. The diagnosis of invasion depth for CRCs using the lateral split-view of a CT enema image, in particular, could play an important diagnostic role for CRCs located on a colorectal fold. The results of the present study show that the measurement of the width of a colorectal - fold lateral contour using a lateral split-view CT enema image can be useful to differentiate intramucosal / T1a-stage CRCs from T1b-stage / more deeply invading CRCs with high diagnostic accuracy.

This study had some limitations. First, there are discrepancies in the pathological diagnosis of colorectal neoplasia between the West and Japan. Namely, intramucosal cancer diagnosed by Japanese pathologists is diagnosed as high-grade dysplasia by the West pathologists. However, this discrepancy does not matter much when deciding on a treatment strategy. Second, the small number of subjects and the fact that this was a single-center retrospective study are also limitations. Further multicenter prospective studies are needed to confirm our initial findings and determine the diagnostic accuracy for the width of a colorectal - fold lateral contour.

## Conclusions

In the depth diagnosis of CRC located on a colorectal fold, the use of a lateral split-view CT enema image can provide higher diagnostic accuracy for quantitative evaluation of a width of a colorectal - fold lateral contour. This may be a new diagnostic approach for determination of the invasion depth of CRCs located on a colorectal fold.

## Data Availability

All relevant data are within the manuscript and its Supporting Information files.

## List of abbreviations

CRC: colorectal cancer
CTC: computed tomographic colonography
CT enema: computed tomographic air-contrast enema
DCBE: double-contrast barium enema
CS-MPR: cross-sectional multiplanar reconstruction
MDCT: multidetector-row computed tomography
ROC: Receiver operating characteristic
ICC: intraclass correlation coefficient

## Notes

### Competing Interest Statement

The authors have declared no competing interest.

### Funding Statement

The author(s) received no specific funding for this work.

### Author Declarations

Ethics committee/IRB of Kyushu University Hospital gave ethical approval for this work.

